# Implemented Interventions in Preventing Surgical Site Infection in Pediatric Appendicitis Patients: A Scoping Review

**DOI:** 10.1101/2024.05.15.24307418

**Authors:** Nutan B. Hebballi, Krysta Sutyak, Maryam Broussard, Caroline Doughty, Elisa Garcia, Kevin P. Lally, Martin L. Blakely, KuoJen Tsao

**Author notes:** **Corresponding Author:** KuoJen Tsao, MD, 6431 Fannin Street, MSB 5.256 Houston, TX 77030.

## Abstract

**Objective:** Surgical site infections (SSIs), especially deep/organ-space SSIs, are common and serious complications following appendectomy. This review aimed to explore the interventions that have been implemented to reduce the risk of SSIs in pediatric appendicitis patients.

**Methods:** A literature search was performed using PubMed, Cochrane, and Embase databases of studies in English published between January 01, 1973, and April 30, 2023. Studies on pediatric patients (≤ 18 years) with appendicitis that described any interventions aimed at reducing SSIs and reported SSIs as an outcome were included.

**Results:** A total of 56 studies were included in the final scoping review. The interventions included antibiotic stewardship, clinical practice guidelines/pathways, different surgical approaches, timing of appendectomy, irrigation or lavage, use of peritoneal drains, timing of wound closure and management, parenteral nutrition, pain management, and outpatient management.

**Conclusion:** A wide variety of interventions have been studied in pediatric appendicitis patients to reduce the SSI rates. Very few publications have studied low-cost, widely available intraoperative interventions to reduce deep/organ-space SSIs.

## Introduction

Acute appendicitis is the most common gastrointestinal disorder affecting children requiring urgent surgical treatment in the United States (US)^1^. Approximately 80,000 children undergo appendectomy annually, and the average cost of surgical care is estimated to be $9,000 per patient^2^. Approximately 30% of acute appendicitis patients present with perforated appendicitis, ranging from 20% to 74%^3,4^. The rate of perforated appendicitis is higher in younger patients^3,4^. In children, perforated appendicitis is associated with post-surgical complications such as wound infections, intra-abdominal abscesses, emergency department visits, hospital readmissions, and extended hospital stay^4,5^.

Surgical site infections (SSIs) are serious postoperative complications that affect 2% of surgical procedures, with varying rates depending upon the surgery type^6^. Typically, surgical wounds are categorized as clean, clean/contaminated, contaminated, and dirty depending upon the bacterial load, and this classification can be used as a predictor of the incidence of SSI for a given surgery^7^. SSIs are the most common complication in children following surgery, resulting in increased morbidity, mortality, additional procedures, longer length of hospital stay, and significant healthcare costs and burdens despite prevention strategies ^8–10^. In pediatric patients, intra-abdominal abscess (also called deep/organ-space surgical site infection) occurs in 10-25% of the patients^11^.

Many strategies, guidelines, and pathways have been proposed to reduce the risk of developing SSIs in pediatric appendicitis and other conditions. These efforts can be aided by implementing a care bundle^12^, utilizing a combination of practices such as antiseptics and prophylactic antibiotics, intraoperative interventions, and postoperative care details^13^. Our baseline impression is that few published studies have focused on widely available, low-cost, intraoperative interventions to reduce deep/organ-space SSIs with perforated appendicitis in children. Most publications likely focus on different antibiotic interventions. In this scoping review, we present the current evidence regarding interventions that have been studied to reduce SSIs in children following appendectomy.

## Methods

### Search strategy

The electronic search was conducted using PubMed, Cochrane, and Embase databases for articles published between January 01, 1973, and April 30, 2023. The inclusion criteria were randomized controlled trials (RCT), observational cohort studies, and interventional (pre-post) studies; pediatric appendicitis patients (≤ 18 years); studies that reported SSI as an outcome; and English language. Studies that described SSI as a wound complication, wound infection, postoperative infection, intra-abdominal abscess, or organ space infection were included. We did not distinguish the type of SSI, such as superficial, deep, organ/space infection, or intra-abdominal abscess. Exclusion criteria were case reports, case series, narrative and systematic reviews, unavailability of full-text, non-English language, microbiological reporting of SSI, and other concurrent surgery performed with appendectomy procedure. Search terms included: “appendicitis”, “appendicitis management”, appendectomy”, “intervention”, “ prevent*”, “surgical site infection”, “SSI”, “wound infection”, “postoperative complication”, “post-operative complication”.

### Study selection

Study selection was completed in two stages. In the first stage, four authors (NH, MB, CD, EG) were divided into two groups, and the titles and abstracts were screened based on the predefined eligibility criteria. In the second stage, articles that appeared pertinent and those with insufficient evidence were included for full-text review and were reviewed by four authors (NH, MB, CD, KS). Discrepancies were resolved through discussions.

### Data extraction and quality assessment

Data from included studies were retrieved and stored in the institutional REDCap^14^, a password-protected secured web database. Extracted data included first author, year of publication, country of publication, study type (single-center or multi-center), study design, intervention details, sample size, whether the SSI was listed as the primary or the secondary outcome, results of SSI and the associated p-values. Based on the interventions described in the study, they were further categorized into subsections that included antibiotic stewardship, clinical practice guidelines/pathways, different surgical approaches, timing of the appendectomy procedure, irrigation or lavage, use of peritoneal drains, timing of wound closure and management, parenteral nutrition, pain management using ketorolac and outpatient management of patients post appendectomy.

## Results

### Search Results

We retrieved 8178 articles from the three database searches, and after removing duplicate records, 6134 articles were screened for titles and abstracts. After this screening, 99 articles were retrieved for full-text reviews, of which six were not found, and 37 were excluded for multiple reasons (Figure 1). Finally, 56 articles met the inclusion criteria and were included in the qualitative analysis (Figure 1).

**Figure 1:**
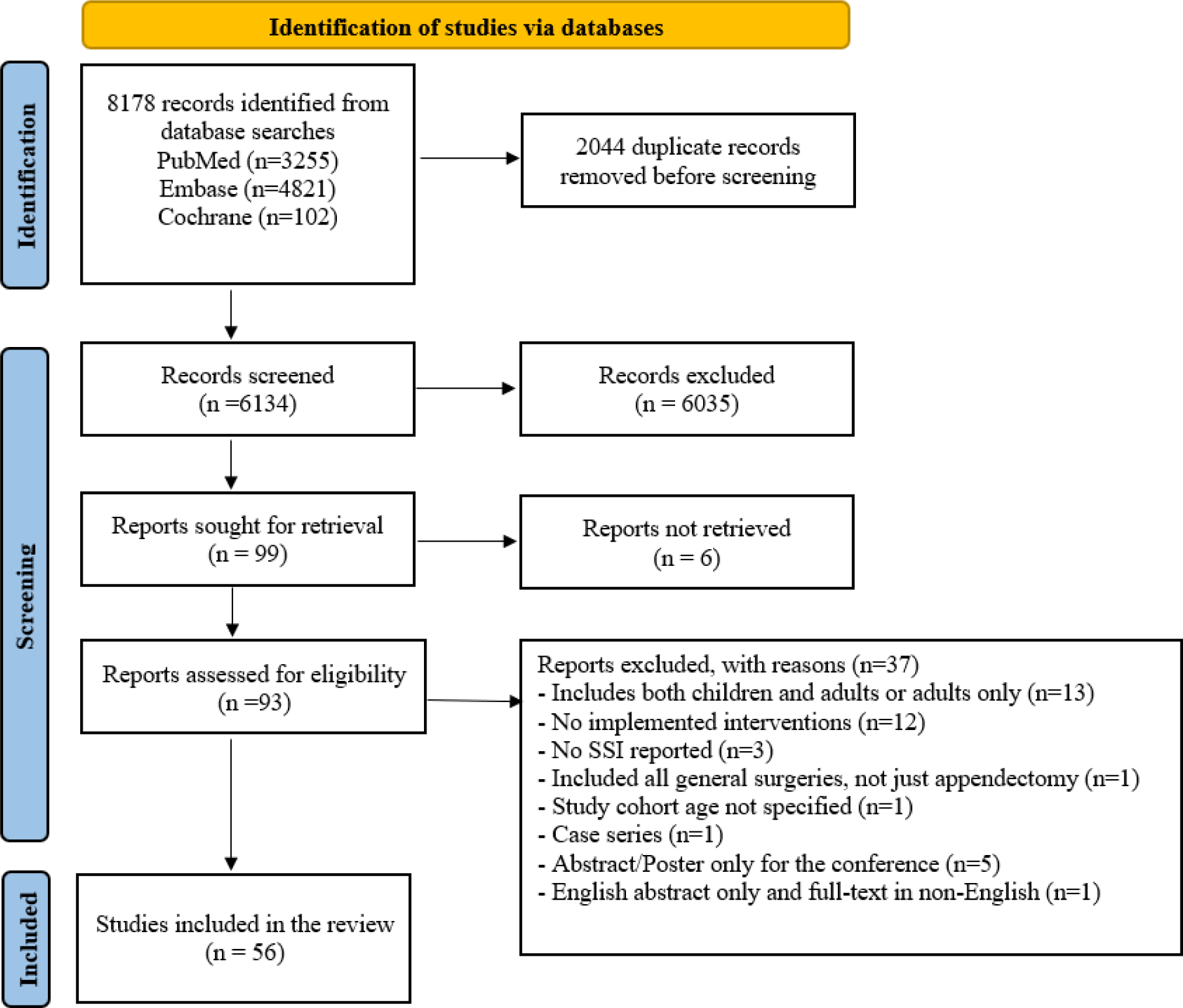
PRISMA flowchart.

### Characteristics of included studies

The main characteristics of all 56 articles are presented in Table 1. Thirty-three studies were performed in the US, 5 studies in Japan, 4 studies in the United Kingdom, 4 studies in Turkey, 2 studies each in Hong Kong and South Africa, and one study each in Australia, Canada, France, Iran, South Korea and Sweden (Table 1). Forty-five studies were single-center, and the remaining 11 were multi-center (Table 1). Regarding study design, 13 were randomized control trials, 5 were prospective cohort studies, 27 were retrospective cohort studies, and 11 were pre-post studies (Table 1). There were 18 studies on antibiotic stewardship, 12 studies each on clinical practice guidelines/pathways and surgical approach, 4 studies on the timing of appendectomy procedure, 3 studies on irrigation or lavage, two studies each on the use of peritoneal drains and timing of wound management and closure and one studies each on parenteral nutrition, pain management using ketorolac and outpatient management after appendectomy (Table 1).

**Table 1:** Characteristics of included studies.

| Study characteristics |  | n (%) |
| --- | --- | --- |
| Design | Randomized controlled trials | 13 (23.2) |
|  | Prospective cohort study | 5 (8.9) |
|  | Retrospective cohort study | 27 (48.2) |
|  | Pre-post study | 11 (19.6) |
| Country | United States | 33 (58.9) |
|  | Japan | 5 (8.9) |
|  | United Kingdom | 4 (7.1) |
|  | Turkey | 4 (7.1) |
|  | Hong Kong | 2 (3.6) |
|  | South Africa | 2 (3.6) |
|  | Australia | 1(1.8) |
|  | Canada | 1(1.8) |
|  | France | 1(1.8) |
|  | Iran | 1(1.8) |
|  | South Korea | 1(1.8) |
|  | Sweden | 1(1.8) |
| Type | Single center | 45 (80.4) |
|  | Multi center | 11 (19.6) |
| Interventions | Antibiotic stewardship | 18 (32.1) |
|  | Clinical practice guidelines/pathways | 12 (21.4) |
|  | Surgical approach | 12 (21.4) |
|  | Irrigation or lavage | 3 (5.4) |
|  | Use of peritoneal drains | 2 (3.6) |
|  | Timing of appendectomy | 4 (7.1) |
|  | Timing of wound closure and management | 2 (3.6) |
|  | Outpatient management after appendectomy | 1 (1.8) |
|  | Parenteral nutrition | 1 (1.8) |
|  | Pain Management | 1 (1.8) |

### Results of individual studies and description of interventions

Evidence from each of the included studies is presented narratively below and in the evidence table (Table 2) according to the intervention types.

**Table 2:**
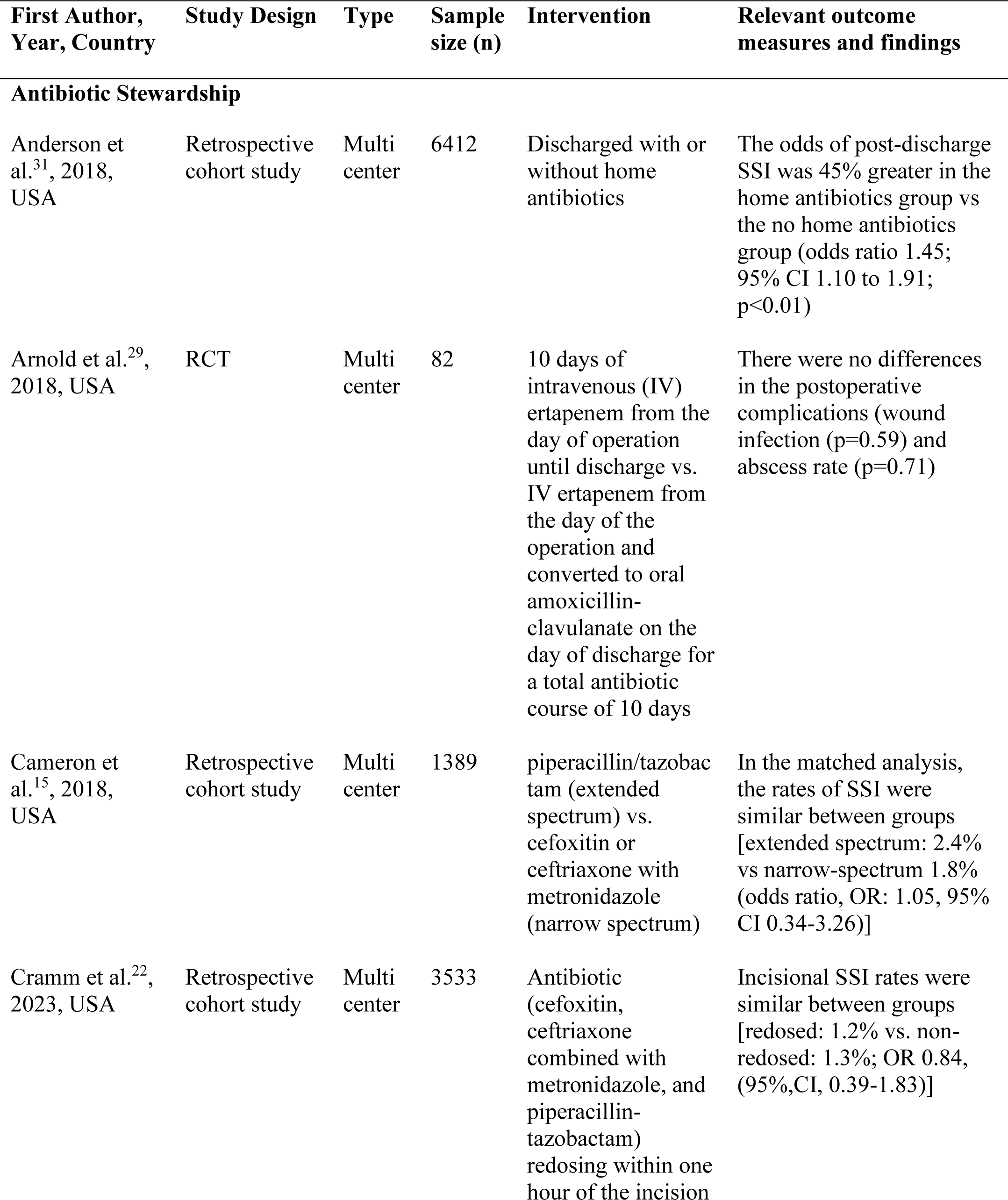
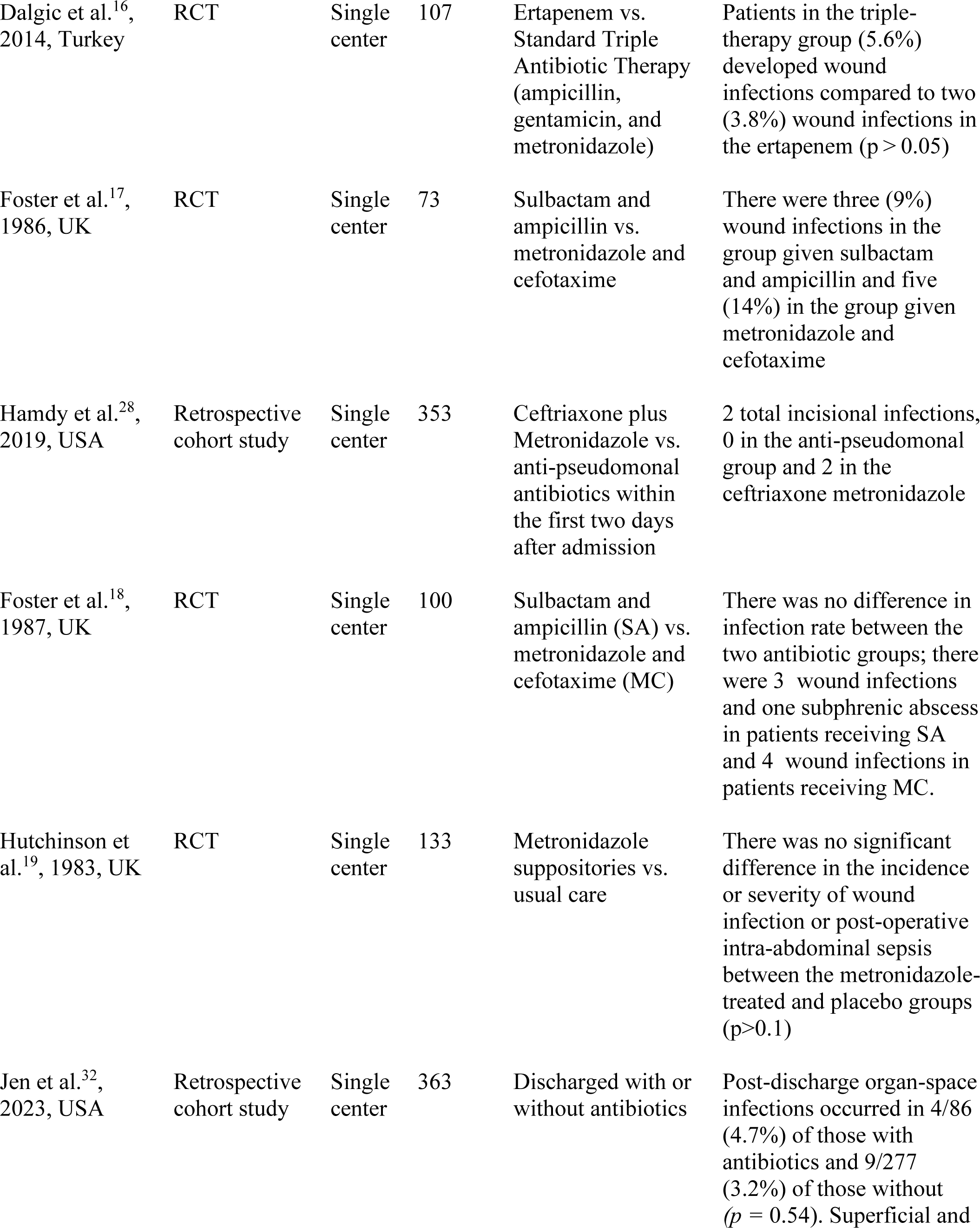
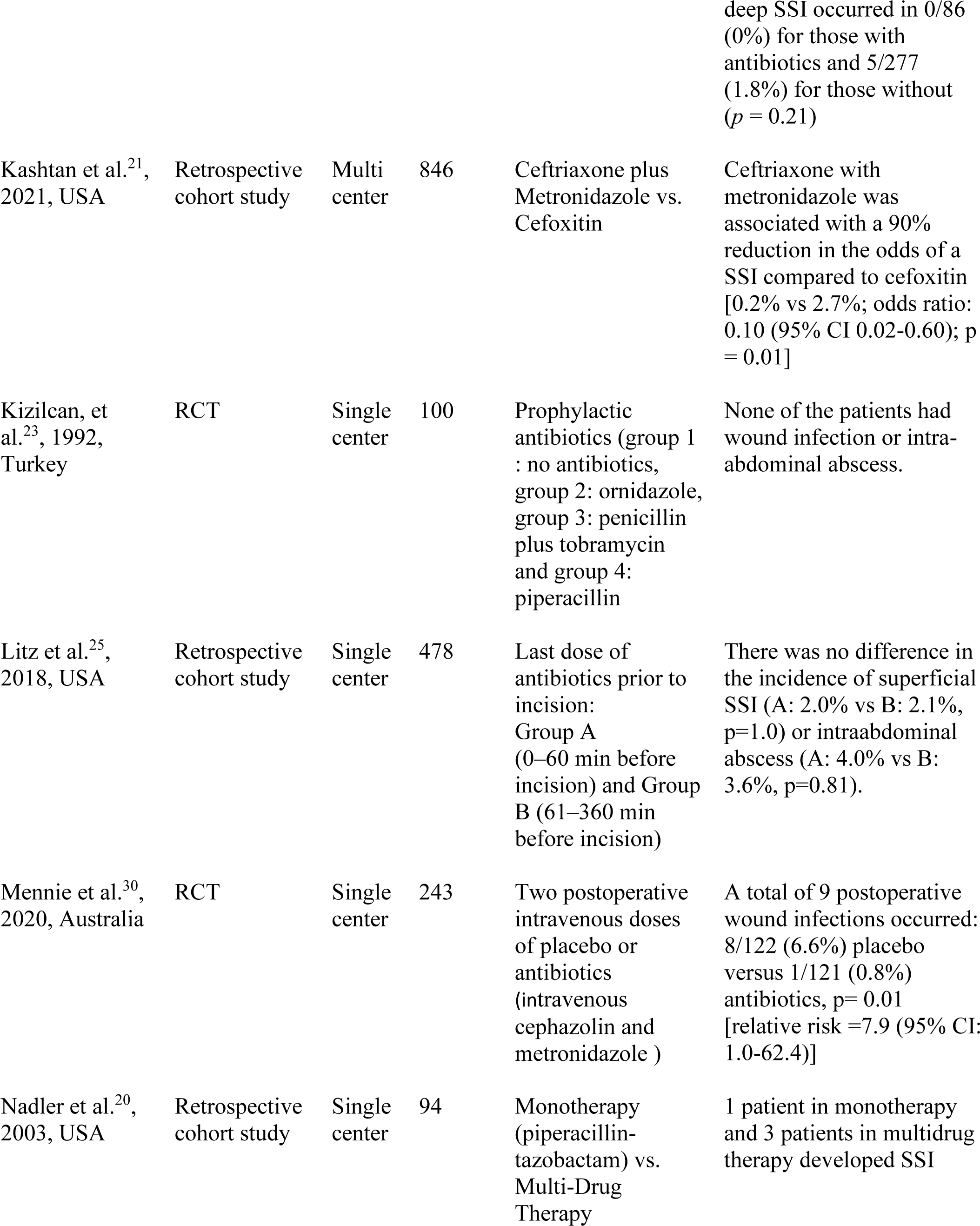
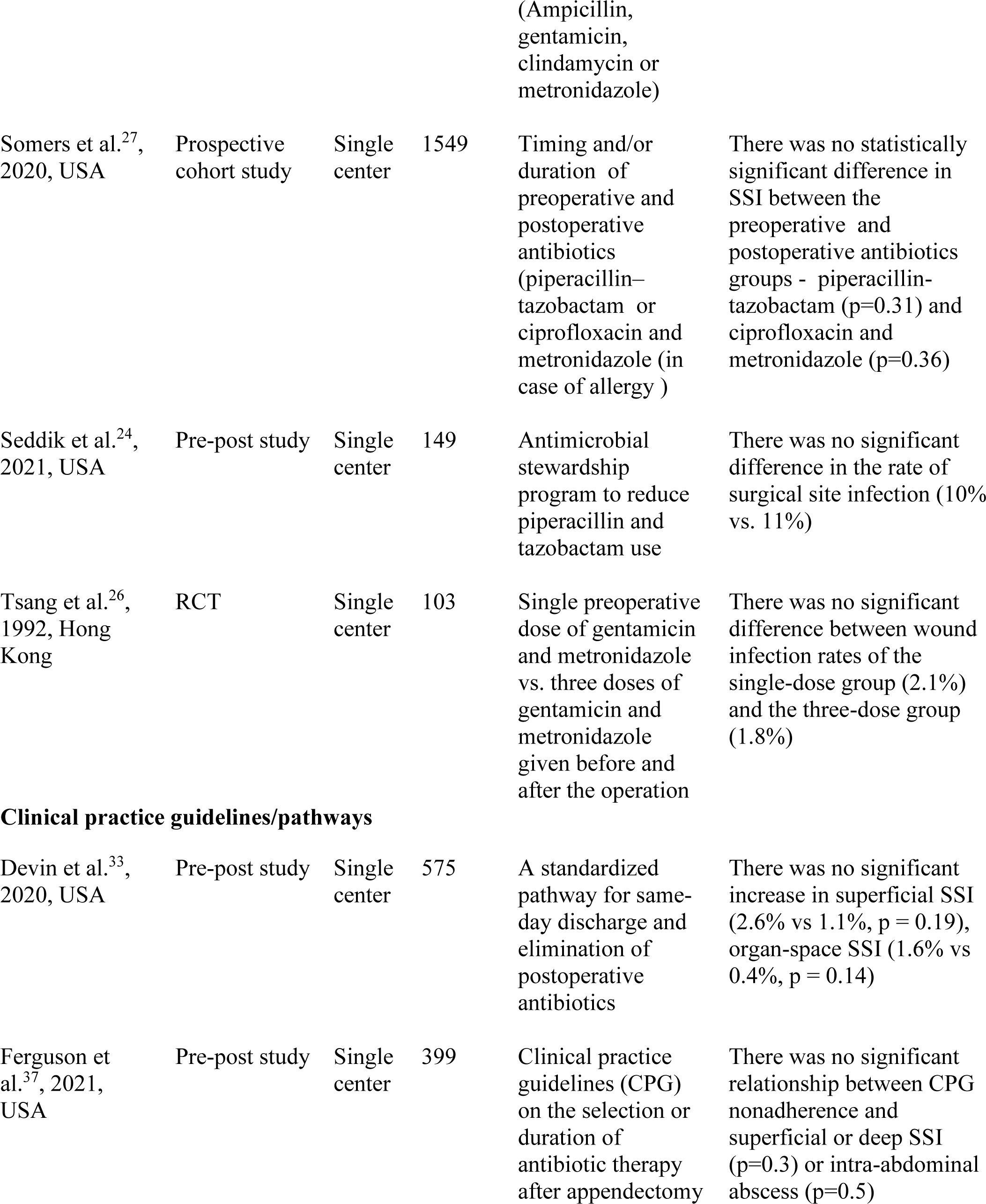
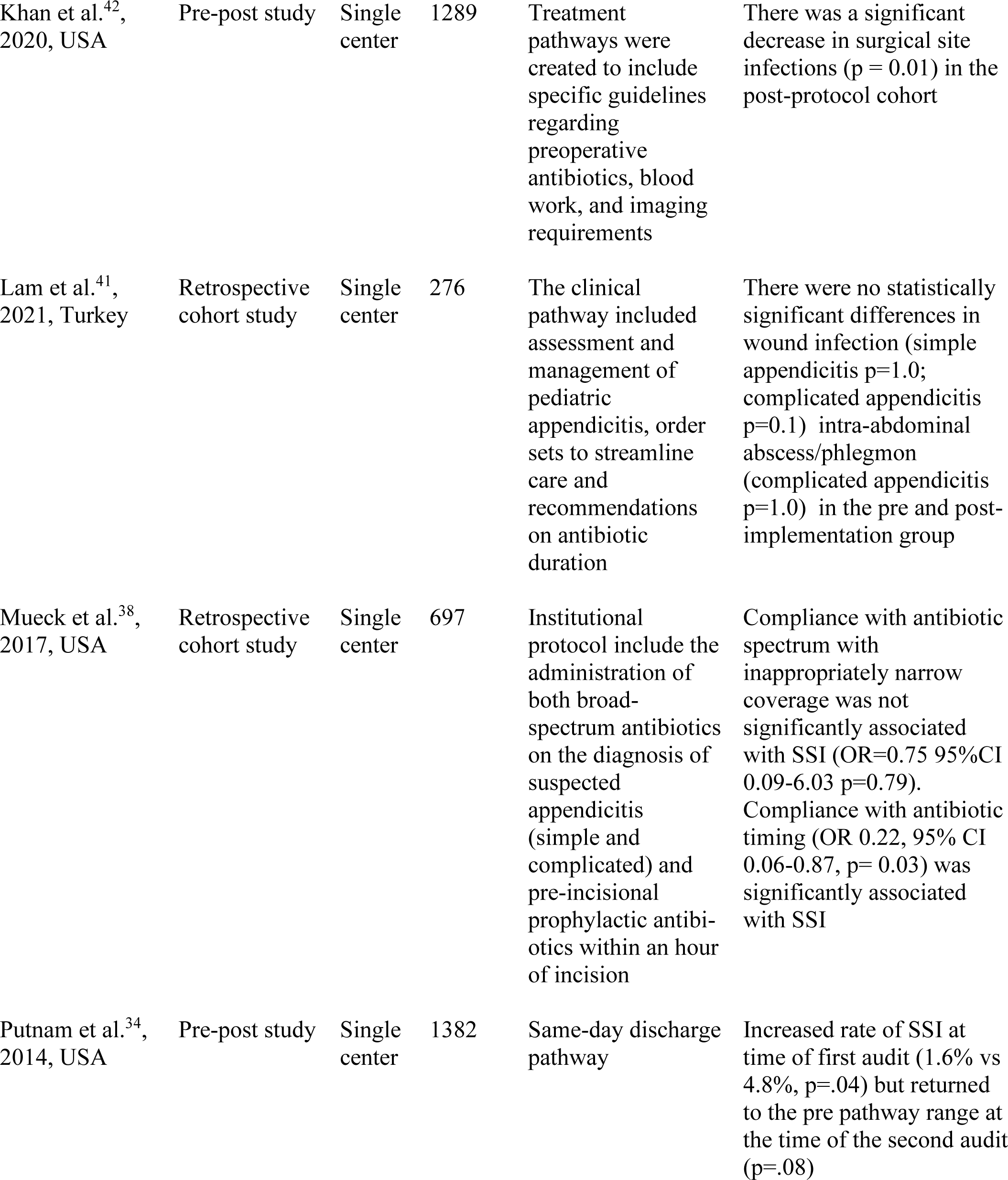
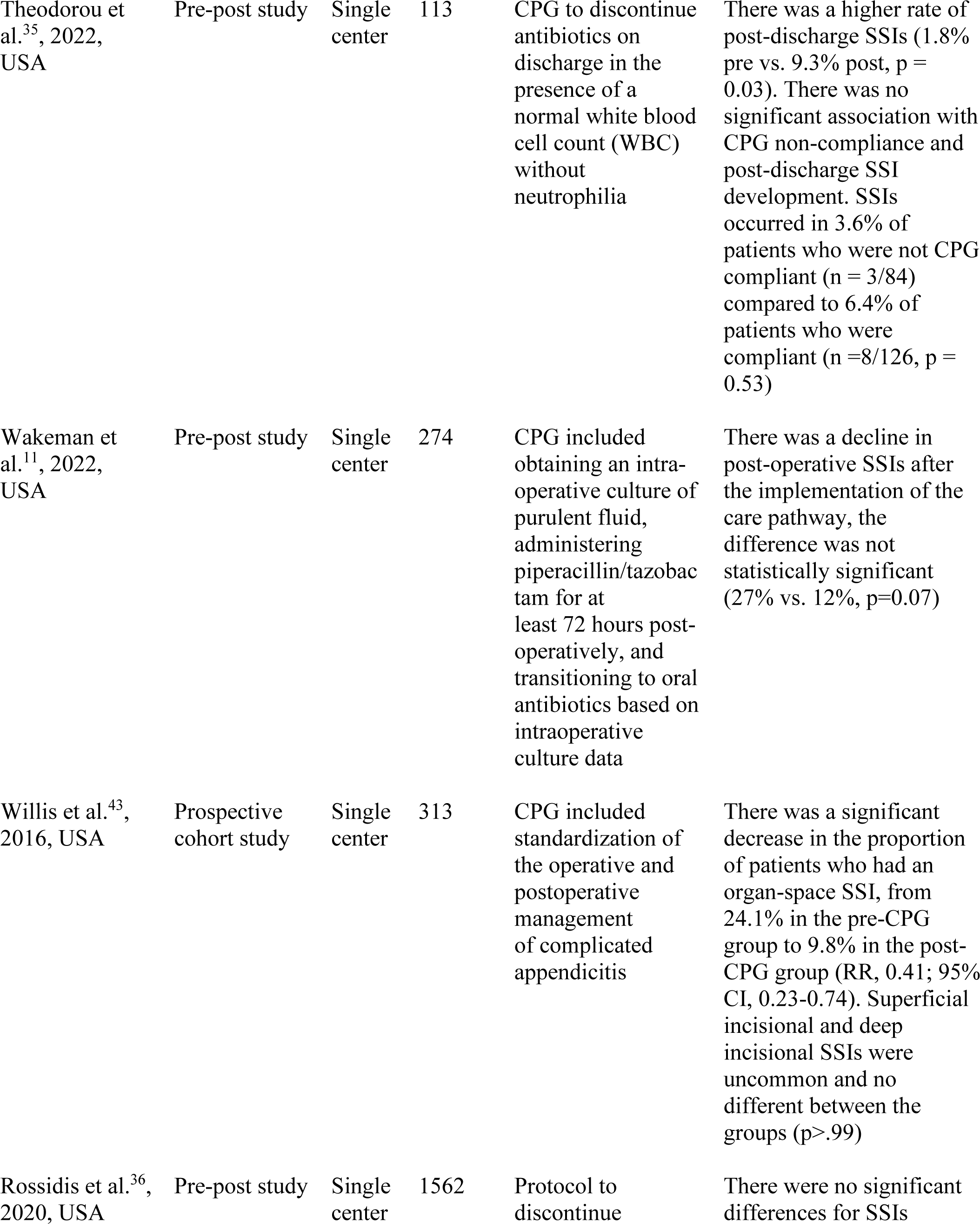
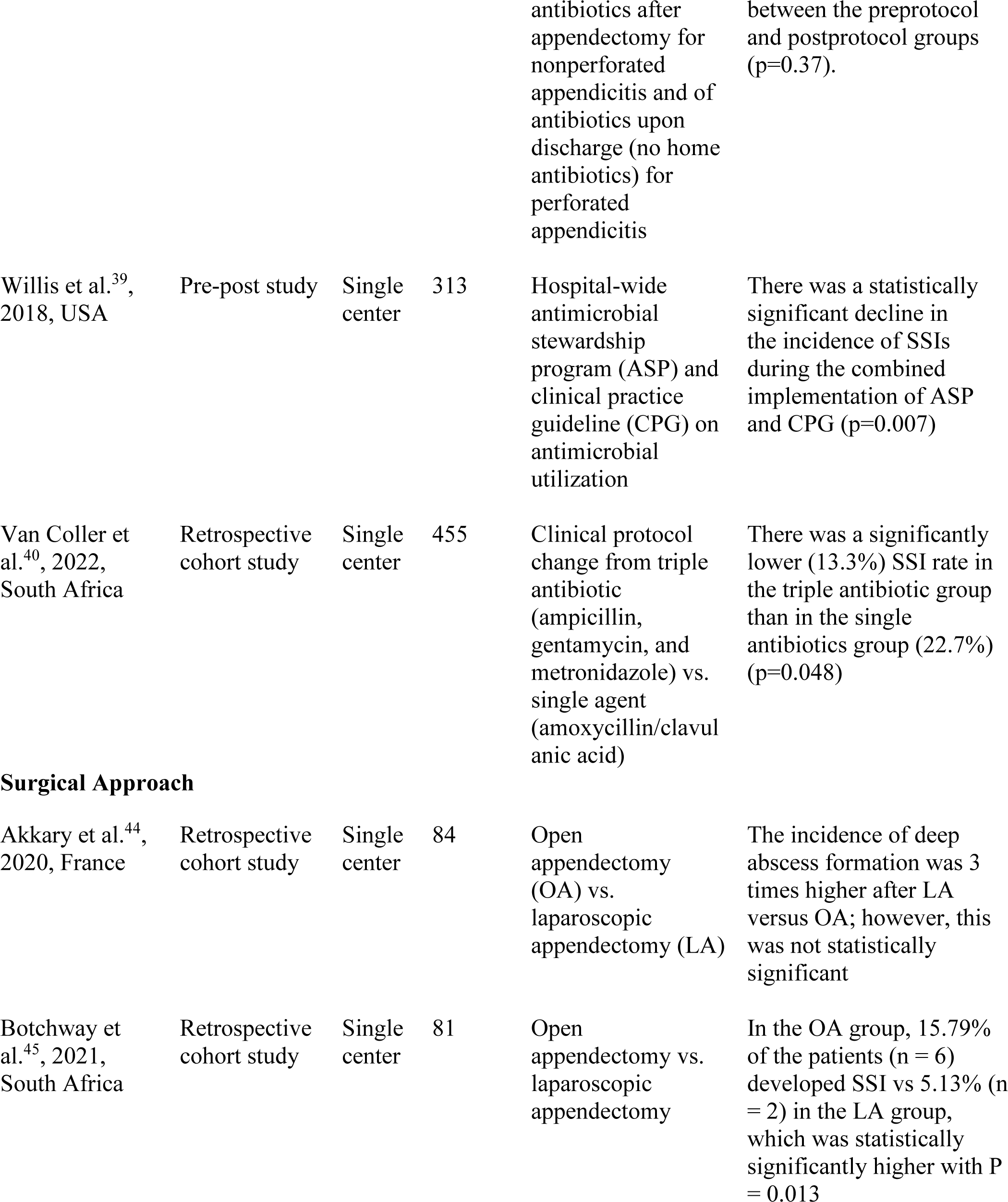
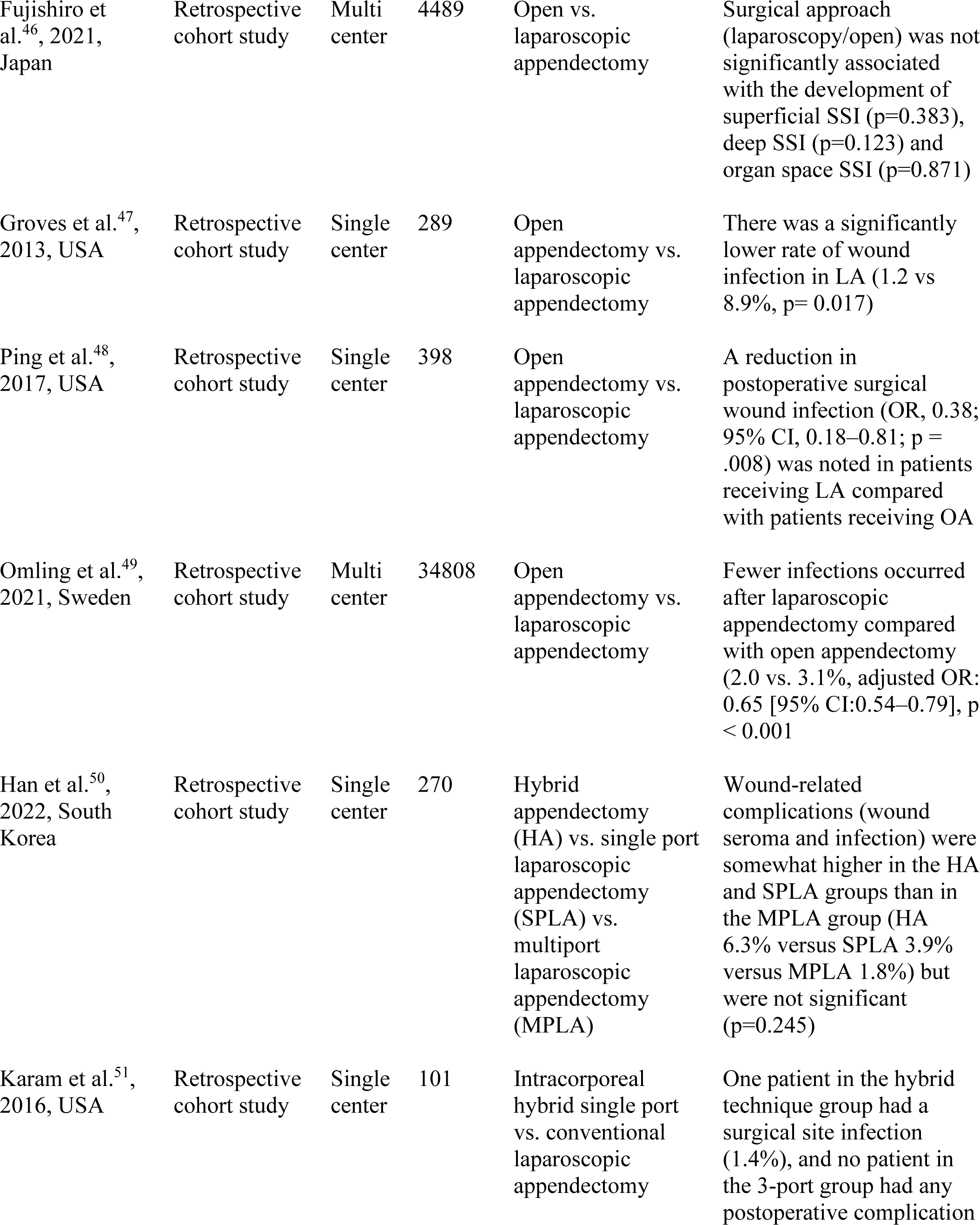
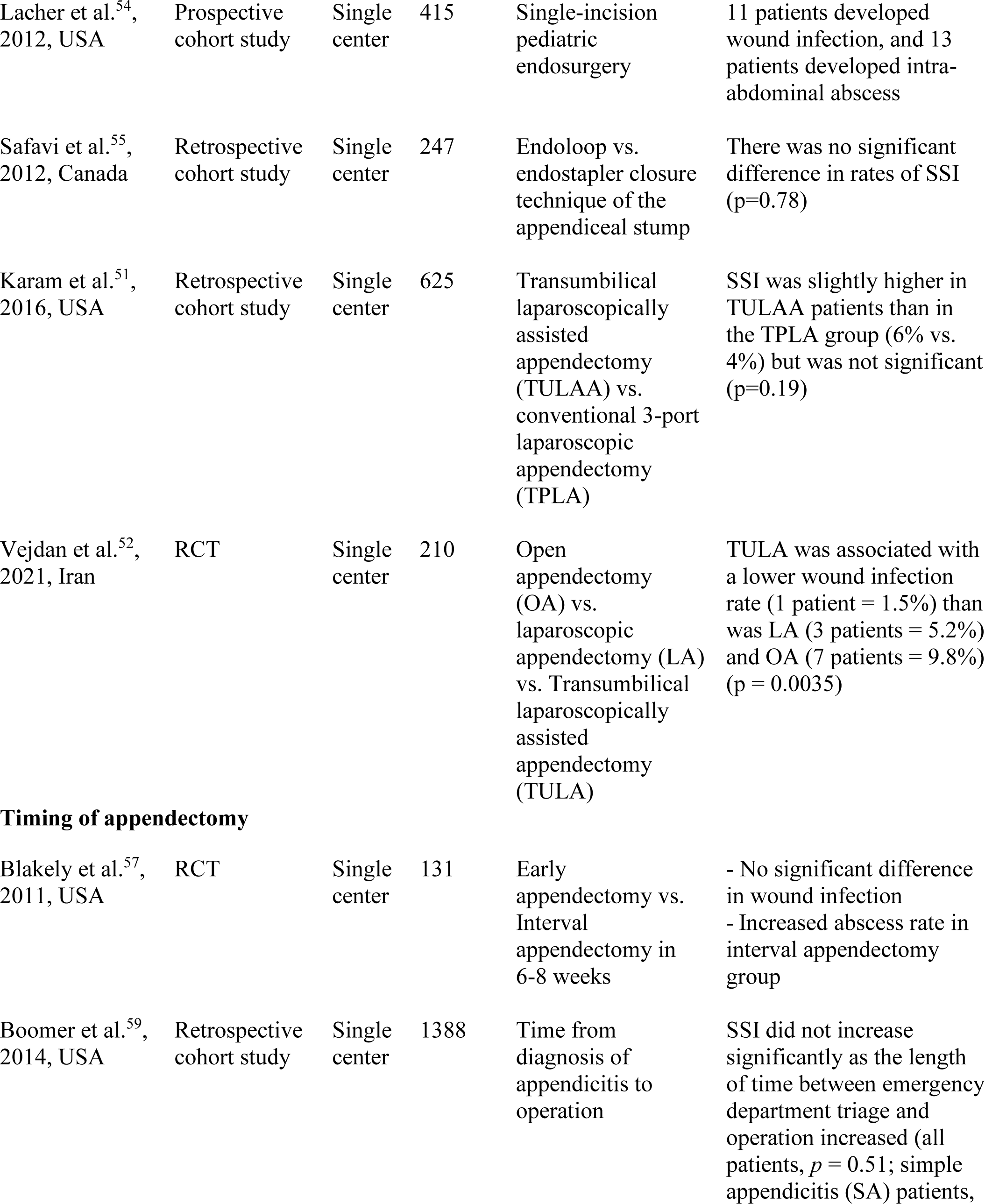
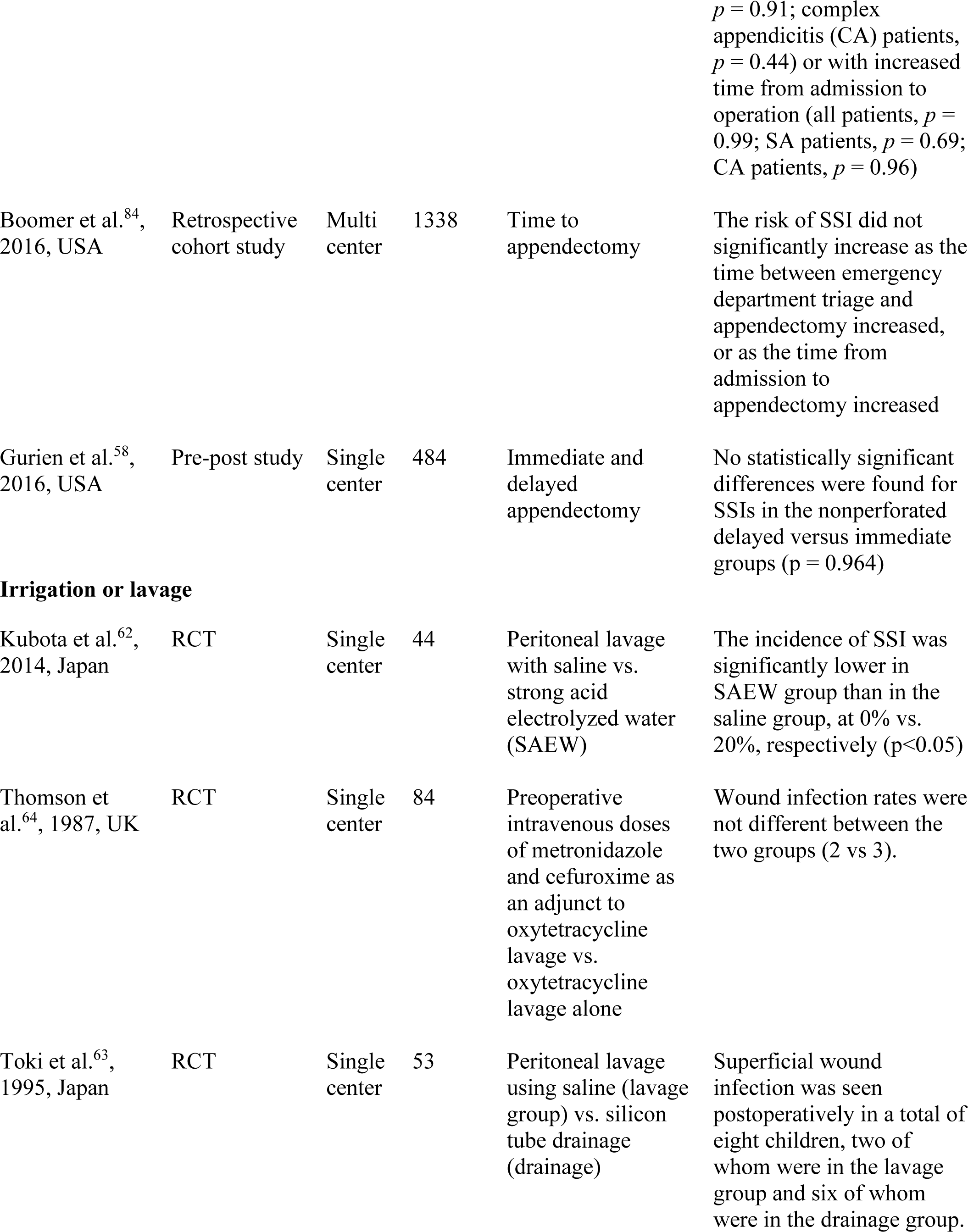
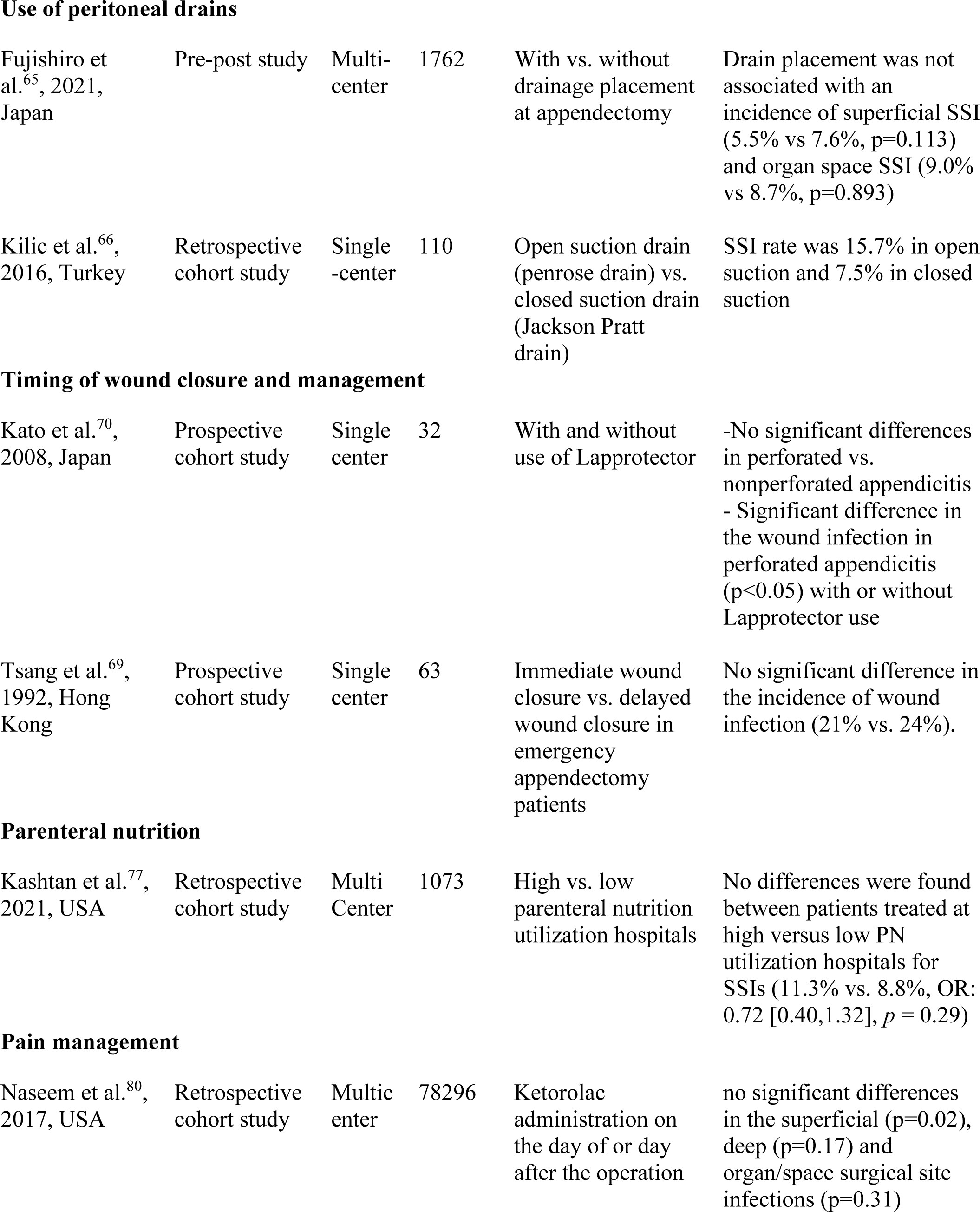
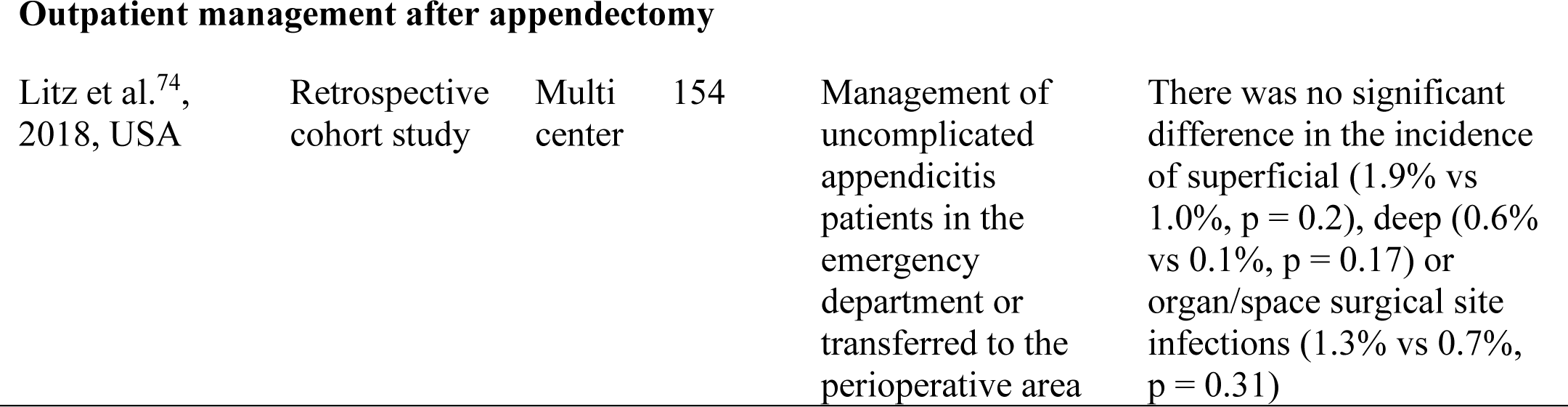
Evidence table of included studies (n=56)

### Antibiotic stewardship

There is considerable variation in antibiotic therapy regarding the choice, duration, timing, route of administration, and discharge of patients at home with or without antibiotics for acute appendicitis management. Seven studies^15–21^ used a combination of antibiotic regimens to reduce the occurrence of SSIs following the appendectomy procedure. Cameron et al. compared the effectiveness of piperacillin/tazobactam (extended-spectrum) vs. cefoxitin or ceftriaxone with metronidazole (narrow spectrum) and found the SSI rates to be similar^15^. Likewise, when ertapenem was compared to the standard triple antibiotic therapy (ampicillin, gentamicin, and metronidazole), Dalgic et al. found no differences in SSIs^16^. Foster et al. compared the effectiveness of sulbactam and ampicillin vs. metronidazole and cefotaxime; they showed no differences in SSIs^17,18^. Hutchinson et al. showed no significant difference in the incidence or severity of wound infection or post-operative intra-abdominal sepsis between the metronidazole-treated and placebo groups^19^. A combination of ceftriaxone and metronidazole was superior in reducing SSIs compared to cefoxitin alone, according to Kashtan et al.^21^.

Antibiotics were administered at different points in time during the patient’s care in the hospital and at discharge. Cramm et al. found no significant difference in the SSI rates after redosing cefoxitin, ceftriaxone combined with metronidazole, and piperacillin-tazobactam within one hour of the appendectomy procedure^22^. None of the patients had wound infection or intra-abdominal abscess with or without administration of ornidazole, penicillin plus tobramycin, and piperacillin prophylactically, as reported by Kizilcan et al. ^23^. In another study by Seddik et al., no significant differences were noted in SSI rates after reducing piperacillin and tazobactam use^24^. No differences in SSI or intra-abdominal abscesses were reported when antibiotics were administered within one hour before incision, as noted by Litz et al.^25^. Similar observations were made by Tsang et al. when patients were administered either a single preoperative dose of gentamicin and metronidazole or three doses of gentamicin and metronidazole given before and after the operation^26^. Somers et al. noted that the timing and duration of preoperative and postoperative antibiotics (piperacillin-tazobactam or ciprofloxacin and metronidazole (in allergy) did not impact SSI rates^27^. Moreover, administration of a combination of ceftriaxone and metronidazole vs. anti-pseudomonal antibiotics within the first two days after admission or 10 days of intravenous ertapenem alone or converted to oral amoxicillin-clavulanate did not affect the SSI rates^28,29^. However, two studies by Mennie et al. and Nadler et al. noted that two doses of intravenous cephazolin and metronidazole reduced the occurrence of wound infection significantly ^30^ and using a multi-drug therapy of ampicillin, gentamicin, clindamycin, or metronidazole increased the occurrence of SSIs when compared to monotherapy (piperacillin-tazobactam)^20^. Anderson et al. reported that patients discharged home had significantly higher SSIs than patients without antibiotics at discharge^31^. In contrast, post-discharge antibiotics did prevent the development of superficial, deep, or organ space infections in post-appendectomy patients, according to Jen et al.^32^

### Clinical practice guidelines/pathways

Various clinical practice guidelines (CPG) or clinical pathways were studied to reduce SSI following appendectomy. Two studies examined the effect of same-day discharge CPG, and no significant reduction in SSIs was observed before or after implementation ^33,34^. However, Putnam et al. found a significant reduction in SSI during their first audit but returned to their usual rates^34^. Two studies that utilized CPGs to discontinue antibiotics had conflicting results. Theodorou et al. found higher post-discharge SSI rates; however, this increase in SSIs was not affected by CPG compliance^35^. In contrast, no significant differences in SSIs between the pre-protocol and post-protocol groups were reported by Rossidis et al.^36^. While the selection of preoperative/postoperative antibiotics varied, its duration did not significantly affect SSI rates^11,37,38^. However, the timing of antibiotics administration was significantly associated with SSIs^38^. Two studies that focused on implementing a combination of hospital-wide antimicrobial stewardship programs and CPG on antimicrobial utilization and changing the triple antibiotic (ampicillin, gentamycin, and metronidazole) to the single agent (amoxicillin/clavulanic acid) demonstrated a significant reduction in SSI rates^39,40^. In three studies that focused their CPGs on the assessment and management of appendicitis preoperatively and postoperatively, including blood work, imaging requirements, and streamlining of care orders sets ^41–43^, only Khan et al. reported a significant reduction in SSI rates in the post CPG implementation cohort^42^.

### Surgical approach

In pediatric patients, two operative modalities, namely open appendectomy and laparoscopic appendectomy, have been widely used for the management of appendicitis. Of the six studies^44–49^ that compared the SSI rates between open appendectomy and laparoscopic appendectomy, four of them found laparoscopic appendectomy had a significantly lower risk of developing SSIs compared to open appendectomy^45,47–49^. In contrast, two studies^44,46^ found no statistically significant association between the operative techniques (open/laparoscopic appendectomy) and SSIs.

Other surgical modalities, such as single-port, multi-port laparoscopic appendectomy, and transumbilical laparoscopic assisted appendectomy, were also studied^50–52^. Han et al. compared the hybrid appendectomy (HA), single port laparoscopic appendectomy (SPLA), and multiport laparoscopic appendectomy (MPLA). They found that wound-related complications such as wound infection and wound seroma were higher in the HA and SPLA groups than in the MPLA group^50^. However, it did not meet statistical significance (p=0.245)^50^. Similarly, one patient in the HA group has SSI, while no patients developed SSIs in a study that compared the intracorporeal hybrid single port vs. conventional laparoscopic appendectomy^53^. In a feasibility study of single-incision pediatric endosurgery for treating appendicitis, Lacher et al. observed that 11 patients and 13 patients developed wound infection and intra-abdominal abscesses, respectively^54^. Furthermore, two studies found a conflicting result for SSI rates when the transumbilical laparoscopically assisted appendectomy (TULA) was compared with conventional 3-port laparoscopic (TPLA), laparoscopic appendectomy, and open appendectomy. Karam et al. found no statistically significant (p=0.19)^51^, but Vejdan et al. demonstrated that TULA was associated with lower wound infection (p=0.0035)^52^. Lastly, in another study, there was no significant difference in rates of SSI when the endoloop versus endostapler technique was used for the closure of the appendiceal stump^55^.

### Timing of appendectomy

Given the emergent nature of acute appendicitis in children, urgent appendectomy is the recognized course of treatment in the United States^56^. However, a few studies assessed the relationship between the effect of time on appendectomy and the risk of developing SSIs in children. A study by Blakely et al. compared early appendectomy with interval appendectomy after 6 to 8 weeks and found that patients treated with interval appendectomy were significantly more likely to develop an intra-abdominal abscess (p=0.02) with no significant differences for wound infection (p=0.91)^57^. Similarly, Gurien et al. found no significant differences in SSI in the nonperforated delayed vs. immediate group (p=0.96)^58^. Additionally, the time to appendectomy and time from diagnosis of appendicitis to the appendectomy operation did not increase the risk of developing SSI in simple and complicated appendicitis^59,60^.

### Irrigation or lavage

To minimize the risk of developing SSI, irrigation has been investigated as a technique to remove residual peritoneal abscess residue in perforated appendicitis. However, the effectiveness of peritoneal irrigation or lavage has been debatable^61^. In our review, we found mixed results, where the incidence of SSI was significantly lower in patients irrigated with strong acid-electrolyzed water (0% vs. 20%, respectively (p<0.05) compared to saline alone^62^ and children with perforated appendicitis who underwent peritoneal lavage with saline had a lower occurrence of superficial wound infection as compared to those who underwent appendectomy with silicon tube drainage^63^. On the contrary, the rate of wound infection did not differ between the group of pediatric patients who were treated with preoperative intravenous doses of metronidazole and cefuroxime as an adjunct to oxytetracycline lavage vs. oxytetracycline lavage alone in non-perforated appendicitis^64^.

### Use of peritoneal drains

The use of a peritoneal drain in complicated appendicitis patients is controversial, given that they pose a significant risk of developing postoperative complications such as wound infection or intra-abdominal abscess^47^. However, in a study conducted by Fujushiro et al., drain placement was not associated with an incidence of superficial SSI (5.5% vs. 7.6%, p=0.113) and organ space SSI (9.0% vs. 8.7%, p=0.893) in pediatric complicated appendicitis patients^65^. In another study, 15.7% of the 70 patients and 7.5% of the 40 patients developed SSI when treated with Penrose drain and Jackson Pratt drain, respectively, in pediatric perforated appendicitis patients^66^.

### Timing of wound closure and management

Wound infection is a common complication in children with perforated or gangrenous appendicitis and those who undergo open appendectomy ^67,68^. Tsang et al. assessed the effect of immediate wound closure and delayed wound closure with skin tape in a prospective study of 63 children undergoing emergency appendectomy and found no significant difference in the incidence of wound infection in perforated appendicitis patients (21% vs. 24%)^69^. In another study, Kato et al. used a Lapprotector, a protective film, and a device to safeguard the open appendectomy wound to prevent infection. They did not find any significant differences in patients with perforated and nonperforated appendicitis patients^70^. However, it was significant in protecting against wound infection in patients with perforated appendicitis (p<0.05), suggesting the use of Lapprotector while performing open appendectomy procedures in perforated appendicitis cases^70^.

### Outpatient management after appendectomy

Several studies have demonstrated that it is safe to discharge patients home the same day after undergoing appendectomy without long-term consequences that help save healthcare cost^71–73^. An analysis of 154 institutional patients and 4973 patients from the American College of Surgeons, National Surgical Quality Improvement Program-Pediatric (NSQIP-P) database demonstrated that there were no significant differences in the superficial (p=0.02), deep (p=0.17) and organ/space surgical site infections (p=0.31) when pediatric uncomplicated appendicitis patients were managed either in the emergency department under observation status or were transferred to the perioperative area without the need for an inpatient hospital admission^74^.

### Parenteral nutrition

Studies have shown that there is variability in care and resource utilization in children with complicated appendicitis ^75,76^. One such variation is in the utilization of parenteral nutrition in complicated appendicitis patients, which can be attributable to the surgeon’s perceptions about parenteral nutrition’s utility in postoperative recovery, wound healing, and immune function, especially in children with increased metabolic needs due to sepsis or inflammatory response^76,77^. However, no significant differences were noted between patients treated at high versus low parenteral nutrition utilization hospitals for SSIs (11.3% vs. 8.8%, OR: 0.72 [95%CI: 0.40,1.32], *p* = 0.29) after matching patients on sex, age, race, payor, body mass index and postoperative hospital length of stay based on the 29 hospitals participating in the NSQIP-Pediatric Appendectomy Pilot Collaborative^77^.

### Pain Management

Ketorolac is a well-known non-steroidal anti-inflammatory drug that the FDA has approved for management of acute pain^78^. In adults, ketorolac can cause complications such as increased emergency department visits and a higher rate of readmission when used for pain management after undergoing gastrointestinal surgeries^79^. However, in pediatric patients, administration of ketorolac on the day of or a day after the appendectomy procedure was not associated with readmission with intra-abdominal postoperative infection within 30 days (p=0.14)^80^. Thus, Naseem et al. recommend using ketorolac during the perioperative period in pediatric appendectomy patients based on their analysis of 78 296 pediatric patients data from the pediatric health information system^80^.

## Discussion

Our comprehensive scoping review was aimed at identifying interventions that have been studied to reduce SSIs in children after appendectomy for acute appendicitis. In our over fifty years (1973-2023) review, we found 56 studies conducted to prevent SSIs after appendectomy in children. These SSI reduction interventions included changes in the combination, duration, and timing of antibiotics, implementation or modifications to the clinical practice guidelines/ clinical pathways, use of different types of surgical approaches, timing of appendectomy procedure, irrigation or lavage, use of peritoneal drains, timing of wound closure and management, parenteral nutrition, pain management using ketorolac and outpatient management of patients after appendectomy.

Given the significant toll of SSIs on patients, hospitals, and the US healthcare system, several guidelines for preventing, detecting, and managing SSIs have been published previously^81,82^. In 2016, the American College of Surgeons (ACS) and Surgical Infection Society published SSI prevention and management guidelines, including prehospital, hospital, and post-discharge interventions for abdominal surgery^83^. Our scoping review identified interventions such as antibiotic stewardship, wound protection, closure, and care that were grouped under hospital interventions in the ACS and Surgical Infection Society’s guidelines.

Our scoping review has some limitations. The scoping reviews have inherent limitations of presenting breadth rather than depth of information on a particular topic. However, our research objective was to map out the current evidence in the literature; thus, this methodology was appropriate. For our scoping review, we did not limit to certain types of appendicitis, such as simple, complicated, gangrenous, or perforated appendicitis. Nonetheless, the interventions we described here have been utilized for reducing SSIs regardless of the type of appendicitis. Lastly, we included studies published in English only due to the vast number of included studies. Thus, our results are generalizable to only published articles written in English. Future studies should focus on assessing these interventions’ long-term implementation, compliance, and cost-effectiveness.

Despite these limitations, our scoping review fills an important gap in the existing pediatric surgical literature by organizing a wide range of studied interventions of evidence-based practices that can be adopted to prevent SSIs in pediatric appendicitis patients.

## Conclusion

In pediatric appendicitis patients, a wide variety of evidence-based interventions have been implemented to prevent SSIs following appendectomy. The effectiveness of the studied interventions varied greatly. Most prior interventions focus on antibiotic considerations, and very few intraoperative interventions have been studied appropriately. Despite these interventions, SSI rates remain high in children, necessitating further investigations. Pediatric surgeons should be cognizant of the implications of SSIs on their patient’s health and on the healthcare system, monitor their own practices, and adopt the interventions that are feasible to prevent occurrences of SSIs.

## Data Availability

No data was produced in this scoping review manuscript. All the research studies that were included in this manuscript are available on PubMed, Cochrane, and Embase databases

## References

1. Guthery SL, Hutchings C, Dean JM, Hoff C. National estimates of hospital utilization by children with gastrointestinal disorders: analysis of the 1997 kids’ inpatient database. J Pediatr. 2004;144(5):589–594. doi:10.1016/j.jpeds.2004.02.029

2. Salvi PS, Cowles RA, Oh PS, Solomon DG. Variability in pediatric appendectomy: The association between disposable supply cost and procedure duration. Surgery. 2022;172(2):729–733. 10.1016/j.surg.2022.04.006

3. Halaseh SA, Kostalas M, Kopec CA, Nimer A. Single-Center Retrospective Analysis of Neutrophil, Monocyte, and Platelet to Lymphocyte Ratios as Predictors of Complicated Appendicitis. Cureus. 2022;14(9):e29177. doi:10.7759/cureus.29177

4. Ribeiro AM, Romero I, Pereira CC, et al. Inflammatory parameters as predictive factors for complicated appendicitis: A retrospective cohort study. Ann Med Surg. 2022;74:103266. doi:10.1016/j.amsu.2022.103266

5. Gosain A, Williams RF, Blakely ML. Distinguishing acute from ruptured appendicitis preoperatively in the pediatric patient. Adv Surg. 2010;44:73–85. doi:10.1016/j.yasu.2010.05.021

6. de Lissovoy G, Fraeman K, Hutchins V, Murphy D, Song D, Vaughn BB. Surgical site infection: incidence and impact on hospital utilization and treatment costs. Am J Infect Control. 2009;37(5):387–397. doi:10.1016/j.ajic.2008.12.010

7. Alganabi M, Biouss G, Pierro A. Surgical site infection after open and laparoscopic surgery in children: a systematic review and meta-analysis. Pediatr Surg Int. 2021;37(8):973–981. doi:10.1007/s00383-021-04911-4

8. Bert F, Giacomelli S, Amprino V, et al. The “bundle” approach to reduce the surgical site infection rate. J Eval Clin Pract. 2017;23(3):642–647. doi:10.1111/jep.12694

9. Khoshbin A, So JP, Aleem IS, Stephens D, Matlow AG, Wright JG. Antibiotic Prophylaxis to Prevent Surgical Site Infections in Children: A Prospective Cohort Study. Ann Surg. 2015;262(2):397–402.

10. Owens CD, Stoessel K. Surgical site infections: epidemiology, microbiology and prevention. J Hosp Infect. 2008;70 Suppl 2:3–10. doi:10.1016/S0195-6701(08)60017-1

11. Wakeman D, Livingston MH, Levatino E, et al. Reduction of surgical site infections in pediatric patients with complicated appendicitis: Utilization of antibiotic stewardship principles and quality improvement methodology. J Pediatr Surg. 2022;57(1):63–73. doi:10.1016/j.jpedsurg.2021.09.031

12. Ching PR. Care Bundles in Surgical Site Infection Prevention: A Narrative Review. Curr Infect Dis Rep. Published online 2024. doi:10.1007/s11908-024-00837-9

13. Koumu MI, Jawhari A, Alghamdi SA, Hejazi MS, Alturaif AH, Aldaqal SM. Surgical Site Infection Post-appendectomy in a Tertiary Hospital, Jeddah, Saudi Arabia. Cureus. 2021;13(7):e16187. doi:10.7759/cureus.16187

14. Harris PA, Taylor R, Thielke R, Payne J, Gonzalez N, Conde JG. Research electronic data capture (REDCap)—A metadata-driven methodology and workflow process for providing translational research informatics support. J Biomed Inform. 2009;42(2):377–381. 10.1016/j.jbi.2008.08.010

15. Cameron DB, Melvin P, Graham DA, et al. Extended Versus Narrow-spectrum Antibiotics in the Management of Uncomplicated Appendicitis in Children. Ann Surg. 2018;268(1):186–192. doi:10.1097/SLA.0000000000002349

16. Dalgic N, Karadag C, Bayraktar B, et al. Ertapenem versus Standard Triple Antibiotic Therapy for the Treatment of Perforated Appendicitis in Pediatric Patients: A Prospective Randomized Trial. Eur J Pediatr Surg. 2013;24(05):410–418. doi:10.1055/s-0033-1352524

17. Foster MC, Kapila L, Morris DL, Slack RCB. A Randomized Comparative Study of Sulbactam plus Ampicillin vs, Metronidazole plus Cefotaxime in the Management of Acute Appendicitis in Children. Clin Infect Dis. 1986;8(Supplement_5):S634–S638. doi:10.1093/clinids/8.Supplement_5.S634

18. Foster MC, Morris DL, Legan C, Kapila L, Slack RC. Perioperative prophylaxis with sulbactam and ampicillin compared with metronidazole and cefotaxime in the prevention of wound infection in children undergoing appendectomy. J Pediatr Surg. 1987;22(9 CC-Anaesthesia CC-Child Health CC-Colorectal CC-Wounds):869–872. doi:10.1016/s0022-3468(87)80658-9

19. Hutchinson GH, Patel BG, Doig CM. A double-blind controlled trial of metronidazole suppositories in children undergoing appendicectomy. Curr Med Res Opin. 1983;8(6):441–445. doi:10.1185/03007998309111751

20. Nadler EP, Reblock KK, Ford HR, Gaines BA. Monotherapy versus multi-drug therapy for the treatment of perforated appendicitis in children. Surg Infect (Larchmt). 2003;4(4):327–333. doi:10.1089/109629603322761382

21. Kashtan MA, Graham DA, Melvin P, et al. Ceftriaxone Combined With Metronidazole is Superior to Cefoxitin Alone in the Management of Uncomplicated Appendicitis in Children. Ann Surg. 2021;274(6):e995–e1000. doi:10.1097/SLA.0000000000003704

22. Cramm SL, Chandler NM, Graham DA, et al. Association Between Antibiotic Redosing Before Incision and Risk of Incisional Site Infection in Children With Appendicitis. Ann Surg. 2023;278(4):e863–e869. doi:10.1097/SLA.0000000000005747

23. Kizilcan F, Tanyel FC, Büyükpamukçu N, Hiçsönmez A. The necessity of prophylactic antibiotics in uncomplicated appendicitis during childhood. J Pediatr Surg. 1992;27(5 CC-Child Health CC-Colorectal CC-Wounds):586–588. doi:10.1016/0022-3468(92)90453-e

24. Seddik TB, Rabsatt LA, Mueller C, et al. Reducing Piperacillin and Tazobactam Use for Pediatric Perforated Appendicitis. J Surg Res. 2021;260:141–148. doi:10.1016/j.jss.2020.11.067

25. Litz CN, Asuncion JB, Danielson PD, Chandler NM. Timing of antimicrobial prophylaxis and infectious complications in pediatric patients undergoing appendectomy. J Pediatr Surg. 2018;53(3):449–451. doi:10.1016/j.jpedsurg.2017.05.005

26. Tsang TM, Tam PK, Saing H. Antibiotic prophylaxis in acute non-perforated appendicitis in children: single dose of metronidazole and gentamicin. J R Coll Surg Edinb. 1992;37(2 CC-Child Health CC-Wounds):110–112. https://www.cochranelibrary.com/central/doi/10.1002/central/CN-00085119/full

27. Somers KK, Eastwood D, Liu Y, Arca MJ. Splitting hairs and challenging guidelines: Defining the role of perioperative antibiotics in pediatric appendicitis patients. J Pediatr Surg. 2020;55(3):406–413. doi:10.1016/j.jpedsurg.2019.07.004

28. Hamdy RF, Handy LK, Spyridakis E, et al. Comparative Effectiveness of Ceftriaxone plus Metronidazole versus Anti-Pseudomonal Antibiotics for Perforated Appendicitis in Children. Surg Infect (Larchmt). 2019;20(5):399–405. doi:10.1089/sur.2018.234

29. Arnold MR, Wormer BA, Kao AM, et al. Home intravenous versus oral antibiotics following appendectomy for perforated appendicitis in children: a randomized controlled trial. Pediatr Surg Int. 2018;34(12 CC-Wounds):1257–1268. doi:10.1007/s00383-018-4343-0

30. Mennie N, Panabokke G, Chang A, et al. Are Postoperative Intravenous Antibiotics Indicated After Laparoscopic Appendicectomy for Simple Appendicitis? A Prospective Double-blinded Randomized Controlled Trial. Ann Surg. 2020;272(2):248–252. doi:10.1097/SLA.0000000000003732

31. Anderson TK, Bartz-Kurycki MA, Kawaguchi AL, et al. Home Antibiotics at Discharge for Pediatric Complicated Appendicitis: Friend or Foe? J Am Coll Surg. 2018;227(2):247–254. doi:10.1016/j.jamcollsurg.2018.04.004

32. Jen J, Hwang R, Mattei P. Post-discharge antibiotics do not prevent intra-abdominal abscesses after appendectomy in children. J Pediatr Surg. 2023;58(2):258–262. doi:10.1016/j.jpedsurg.2022.10.024

33. Devin CL, D’Cruz R, Linden AF, et al. Reducing resource utilization for patients with uncomplicated appendicitis through use of same-day discharge and elimination of postoperative antibiotics. J Pediatr Surg. 2020;55(12):2591–2595. doi:10.1016/j.jpedsurg.2020.04.003

34. Putnam LR, Levy SM, Johnson E, et al. Impact of a 24-hour discharge pathway on outcomes of pediatric appendectomy. Surgery. 2014;156(2):455–461. doi:10.1016/j.surg.2014.03.030

35. Theodorou CM, Lee SY, Lawrence Y, Saadai P, Hirose S, Brown EG. The Utility of Discharge Antibiotics in Pediatric Perforated Appendicitis Without Leukocytosis. J Surg Res. 2022;275:48–55. doi:10.1016/j.jss.2022.01.024

36. Rossidis AC, Brown EG, Payton KJ, Mattei P. Implementation of an evidence-based protocol after appendectomy reduces unnecessary antibiotics. J Pediatr Surg. 2020;55(11):2379–2386. doi:10.1016/j.jpedsurg.2020.07.001

37. Ferguson DM, Ferrante AB, Orr HA, et al. Clinical Practice Guideline Nonadherence and Patient Outcomes in Pediatric Appendicitis. J Surg Res. 2021;257:135–141. doi:10.1016/j.jss.2020.07.042

38. Mueck KM, Putnam LR, Anderson KT, Lally KP, Tsao K, Kao LS. Does compliance with antibiotic prophylaxis in pediatric simple appendicitis matter? J Surg Res. 2017;216:1–8.

39. Willis ZI, Duggan EM, Gillon J, Blakely ML, Di Pentima MC. Improvements in Antimicrobial Prescribing and Outcomes in Pediatric Complicated Appendicitis. Pediatr Infect Dis J. 2018;37(5):429–435. doi:10.1097/INF.0000000000001816

40. van Coller R, Arnold M, le Roux H, et al. Amoxycillin/Clavulanic acid monotherapy in complicated paediatric appendicitis: Good enough? J Pediatr Surg. 2022;57(6):1115–1118. doi:10.1016/j.jpedsurg.2022.01.032

41. Lam JY, Beaudry P, Simms BA, Brindle ME. Impact of implementing a fast-track protocol and standardized guideline for the management of pediatric appendicitis. Can J Surg. 2021;64(4):E364–E370. doi:10.1503/cjs.005420

42. Khan S, Siow VS, Lewis A, et al. An Evidence-Based Care Protocol Improves Outcomes and Decreases Cost in Pediatric Appendicitis. J Surg Res. 2020;256:390–396. doi:10.1016/j.jss.2020.05.067

43. Willis ZI, Duggan EM, Bucher BT, et al. Effect of a Clinical Practice Guideline for Pediatric Complicated Appendicitis. JAMA Surg. 2016;151(5):e160194. doi:10.1001/jamasurg.2016.0194

44. Akkary R, Zeidan S, Matta R, Lakis C, Diab N. Pediatric appendectomy in developing countries. Int J Pediatr Adolesc Med. 2020;7(2):70–73. doi:10.1016/j.ijpam.2019.06.006

45. Botchway E, Marcisz L, Schoeman H, Kofi Botchway PP, Mabitsela EM, Tshifularo N. Laparoscopic versus open appendectomy: A retrospective cohort study on the management of acute appendicitis (simple and complicated) in children under 13 years of age. Afr J Paediatr Surg. 2021;18(4):182–186. doi:10.4103/ajps.AJPS_102_20

46. Fujishiro J, Watanabe E, Hirahara N, et al. Laparoscopic Versus Open Appendectomy for Acute Appendicitis in Children: a Nationwide Retrospective Study on Postoperative Outcomes. J Gastrointest Surg. 2021;25(4):1036–1044. doi:10.1007/s11605-020-04544-3

47. Groves LB, Ladd MR, Gallaher JR, et al. Comparing the cost and outcomes of laparoscopic versus open appendectomy for perforated appendicitis in children. Am Surg. 2013;79(9):861–864.

48. Li P, Han Y, Yang Y, et al. Retrospective review of laparoscopic versus open surgery in the treatment of appendiceal abscess in pediatric patients. Medicine (Baltimore). 2017;96(30):e7514. doi:10.1097/MD.0000000000007514

49. Omling E, Salö M, Saluja S, et al. A Nationwide Cohort Study of Outcome after Pediatric Appendicitis. Eur J Pediatr Surg. 2021;31(02):191–198. doi:10.1055/s-0040-1712508

50. Han J, Kim H, Han SH, Kang BM. Hybrid Appendectomy in Pediatric Appendicitis: A Comparative Analysis of Single-Port and Multiport Laparoscopic Appendectomy. J Laparoendosc Adv Surg Tech. 2022;32(3):330–335. doi:10.1089/lap.2021.0625

51. Karam PA, Mohan A, Buta MR, Seifarth FG. Comparison of Transumbilical Laparoscopically Assisted Appendectomy to Conventional Laparoscopic Appendectomy in Children. Surg Laparosc Endosc Percutan Tech. 2016;26(6):508–512. doi:10.1097/SLE.0000000000000334

52. Vejdan SAK, Khosravi M, Amirian Z. Transumbilical laparoscopic-assisted appendectomy as a safe procedure for pediatric uncomplicated appendicitis: a comparison with laparoscopic and open appendectomy in a randomized clinical trial. J Pediatr Endosc Surg. 2021;3(1):39–46. doi:10.1007/s42804-020-00087-1

53. Karam PA, Hiuser A, Magnuson D, Seifarth FGF. Intracorporeal hybrid single port vs conventional laparoscopic appendectomy in children. La Pediatr Medica e Chir. 2016;38(3):89–92. doi:10.4081/pmc.2016.133

54. Lacher M, Muensterer OJ, Yannam GR, et al. Feasibility of Single-Incision Pediatric Endosurgery for Treatment of Appendicitis in 415 Children. J Laparoendosc Adv Surg Tech. 2012;22(6):604–608. doi:10.1089/lap.2012.0107

55. Safavi A, Langer M, Skarsgard E. Endoloop versus endostapler closure of the appendiceal stump in pediatric laparoscopic appendectomy. Can J Surg. 2012;55(1):37–40. doi:10.1503/cjs.023810

56. Warner BW, Kulick RM, Stoops MM, Mehta S, Stephan M, Kotagal UR. An evidenced-based clinical pathway for acute appendicitis decreases hospital duration and cost. J Pediatr Surg. 1998;33(9):1371–1375. doi:10.1016/s0022-3468(98)90010-0

57. Blakely ML, Williams R, Dassinger MS, et al. Early vs Interval Appendectomy for Children With Perforated Appendicitis. Arch Surg. 2011;146(6):660. doi:10.1001/archsurg.2011.6

58. Gurien LA, Wyrick DL, Smith SD, Dassinger MS. Optimal timing of appendectomy in the pediatric population. J Surg Res. 2016;202(1):126–131. doi:10.1016/j.jss.2015.12.045

59. Boomer LA, Cooper JN, Deans KJ, et al. Does delay in appendectomy affect surgical site infection in children with appendicitis? J Pediatr Surg. 2014;49(6):1026–1029. doi:10.1016/j.jpedsurg.2014.01.044

60. Boomer LA, Cooper JN, Anandalwar S, et al. Delaying Appendectomy Does Not Lead to Higher Rates of Surgical Site Infections. Ann Surg. 2016;264(1):164–168. doi:10.1097/SLA.0000000000001396

61. Nataraja R, Panabokke G, Chang A, et al. Does Peritoneal Lavage Influence the Rate of Complications Following Pediatric Laparoscopic Appendicectomy in Children with Complicated Appendicitis? A Prospective Randomized Clinical Trial. J Pediatr Surg. 2019;54. doi:10.1016/j.jpedsurg.2019.08.039

62. Kubota A, Goda T, Tsuru T, et al. Efficacy and safety of strong acid electrolyzed water for peritoneal lavage to prevent surgical site infection in patients with perforated appendicitis. Surg Today. 2015;45(7 CC-Wounds):876–879. doi:10.1007/s00595-014-1050-x

63. Toki A, Ogura K, Horimi T, et al. Peritoneal lavage versus drainage for perforated appendicitis in children. Surg Today. 1995;25(3 CC-Anaesthesia CC-Child Health CC-Colorectal CC-Wounds):207–210. doi:10.1007/BF00311528

64. Thomson SR, Carle G, Reid TM, Davidson AI, Miller SS. Antibiotic prophylaxis in non-perforated appendicitis of childhood: tetracycline lavage compared with peroperative intravenous cefuroxime and metronidazole. J Hosp Infect. 1987;9(2 CC-HS-HANDSRCH CC-Anaesthesia CC-Child Health CC-Colorectal CC-Wounds):158–161. doi:10.1016/0195-6701(87)90054-5

65. Fujishiro J, Fujiogi M, Hirahara N, et al. Abdominal Drainage at Appendectomy for Complicated Appendicitis in Children. Ann Surg. 2021;274(6):e599–e604. doi:10.1097/SLA.0000000000003804

66. Kılıç ŞS, Ekinci S, Karnak İ, Çiftçi AÖ, Tanyel FC, Şenocak ME. Drainage Systems’ Effect on Surgical Site Infection in Children with Perforated Appendicitis. Ann Clin Anal Med. 2016;07(05):591–594. doi:10.4328/JCAM.2865

67. Pettigrew RA. Delayed primary wound closure in gangrenous and perforated appendicitis. Br J Surg. 1981;68(9):635–638. doi:10.1002/bjs.1800680910

68. Ikeda H, Ishimaru Y, Takayasu H, Okamura K, Kisaki Y, Fujino J. Laparoscopic versus open appendectomy in children with uncomplicated and complicated appendicitis. J Pediatr Surg. 2004;39(11):1680–1685. doi:10.1016/j.jpedsurg.2004.07.018

69. Tsang TM, Tam PK, Saing H. Delayed primary wound closure using skin tapes for advanced appendicitis in children. A prospective, controlled study. Arch Surg. 1992;127(4 CC-Anaesthesia CC-Child Health CC-Colorectal CC-Wounds):451–453. doi:10.1001/archsurg.1992.01420040097017

70. Kato Y, Marusasa T, Ichikawa S, Lane GJ, Okazaki T, Yamataka A. Lapprotector^TM^ Use Decreases Incisional Wound Infections in Cases of Perforated Appendicitis: A Prospective Study. Asian J Surg. 2008;31(3):101–103. doi:10.1016/S1015-9584(08)60068-8

71. Farach SM, Danielson PD, Walford NE, Harmel RP, Chandler NM. Same-day Discharge after Appendectomy Results in Cost Savings and Improved Efficiency. Am Surg. 2014;80(8):787–791. doi:10.1177/000313481408000829

72. Aguayo P, Alemayehu H, Desai AA, Fraser JD, St. Peter SD. Initial experience with same day discharge after laparoscopic appendectomy for nonperforated appendicitis. J Surg Res. 2014;190(1):93–97. 10.1016/j.jss.2014.03.012

73. Putnam L, Levy S, Johnson E, et al. Same-Day Discharge for Simple Pediatric Appendicitis: Sustainable Quality Improvement Requires Ongoing Surveillance. J Surg Res. 2014;186(2):615. doi:10.1016/j.jss.2013.11.602

74. Litz CN, Stone L, Alessi R, Walford NE, Danielson PD, Chandler NM. Impact of outpatient management following appendectomy for acute appendicitis: An ACS NSQIP-P analysis. J Pediatr Surg. 2018;53(4):625–628. doi:10.1016/j.jpedsurg.2017.06.023

75. Newman K, Ponsky T, Kittle K, et al. Appendicitis 2000: variability in practice, outcomes, and resource utilization at thirty pediatric hospitals. J Pediatr Surg. 2003;38(3):372–379. doi:10.1053/jpsu.2003.50111

76. Rice-Townsend S, Barnes JN, Hall M, Baxter JL, Rangel SJ. Variation in Practice and Resource Utilization Associated With the Diagnosis and Management of Appendicitis at Freestanding Children’s Hospitals: Implications for Value-Based Comparative Analysis. Ann Surg. 2014;259(6).

77. Kashtan MA, Graham DA, Anandalwar SP, Hills-Dunlap JL, Rangel SJ. Variability, outcomes and cost associated with the use of parenteral nutrition in children with complicated appendicitis: A hospital-level propensity matched analysis. J Pediatr Surg. 2021;56(12):2299–2304. doi:10.1016/j.jpedsurg.2021.03.005

78. Levin RA. Food and Drug Administration.; 2009. https://www.accessdata.fda.gov/drugsatfda_docs/nda/2010/022382Orig1s000MedR.pdf

79. Kotagal M, Hakkarainen TW, Simianu V V, Beck SJ, Alfonso-Cristancho R, Flum DR. Ketorolac Use and Postoperative Complications in Gastrointestinal Surgery. Ann Surg. 2016;263(1):71–75. doi:10.1097/SLA.0000000000001260

80. Naseem H-R, Dorman RM, Ventro G, Rothstein DH, Vali K. Safety of perioperative ketorolac administration in pediatric appendectomy. J Surg Res. 2017;218:232–236. doi:10.1016/j.jss.2017.05.087

81. Mazuski JE, Tessier JM, May AK, et al. The Surgical Infection Society Revised Guidelines on the Management of Intra-Abdominal Infection. Surg Infect (Larchmt). 2017;18(1):1–76. doi:10.1089/sur.2016.261

82. Mangram AJ, Horan TC, Pearson ML, Silver LC, Jarvis WR. Guideline for Prevention of Surgical Site Infection, 1999. Centers for Disease Control and Prevention (CDC) Hospital Infection Control Practices Advisory Committee. Am J Infect Control. 1999;27(2):97–132; quiz 133-134; discussion 96.

83. Ban KA, Minei JP, Laronga C, et al. American College of Surgeons and Surgical Infection Society: Surgical Site Infection Guidelines, 2016 Update. J Am Coll Surg. 2017;224(1):59–74. doi:10.1016/j.jamcollsurg.2016.10.029

84. Boomer LA, Cooper JN, Anandalwar S, et al. Delaying Appendectomy Does Not Lead to Higher Rates of Surgical Site Infections: A Multi-institutional Analysis of Children With Appendicitis. Ann Surg. 2016;264(1):164–168. doi:10.1097/SLA.0000000000001396

